# Prediction of COVID-19 cases during Tokyo’s Olympic and Paralympic Games

**DOI:** 10.1101/2021.04.21.21255676

**Authors:** Yasuharu Tokuda, Toshikazu Kuniya

## Abstract

Tokyo’s Olympic and Paralympic Games set to begin in late July 2021 without spectators from abroad, but vaccine rollout has been slow in Japan compared to other developed countries. In this study, COVID-19 epidemic curve in Tokyo is developed based on weekly reported data from January 23, 2020 until April 16, 2021. The maximum daily number of the infected cases in Tokyo in August 2021 would be 7,991 if the current pace of vaccinations (1/1,000 per day). This daily number is greater than the highest daily cases (2,447) recorded on January 7, 2021. However, if the rollout pace could be doubled (1/500 per day), the peak daily new cases would be 4,470 in August. If it could be quadrupled (1/250 per day), the peak would be noted at 2,128 in July and the highest number in August would be 1,977. If vaccine rollout could not be enhanced, the cancellation might be an acceptable decision, since health is the most precious to our Olympians.

## TEXT

New COVID-19 cases in Japan have increased in recent weeks and the country experiences a “fourth wave” of the infection as it prepares for the Olympic and Paralympic Games, which are set to begin in late July 2021 without spectators from abroad. However, vaccine rollout has been slow in Japan compared to other developed countries. Japan does not produce domestic anti-COVID-19 vaccines and we import and provides mRNA vaccines to people (first healthcare workers, followed by elderly) that started on the February 17, 2021. This is worrisome because there would be a big COVID-19 infection surge in Tokyo during the Games with an overwhelmed local healthcare system and less availability of healthcare workers supporting the Games (1). Moreover, athletes, coaches, judges, and journalists would face greater risk of infection from the surge.

We developed COVID-19 epidemic curve in Tokyo based on weekly reported data from January 23, 2020 until April 16, 2021 (2). A susceptible-exposed-infected-removed compartmental model as in (3) was used for the curve fitting by updating the estimation per week. More precisely, the model equations are as follows.

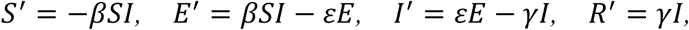

where *S*(*t*), *E*(*t*), *I*(*t*) and *R*(*t*) denote the susceptible, exposed, infected and removed populations at time (day) *t*, respectively. *β* denotes the infection rate and ε and *γ* denote the transition rates from *E* to *I* and *I* to *R*, respectively. As in (3), we set ε = 0.2 and *γ* = 0.1 so that the average period of incubation and infection are 1/ε = 5 (days) and 1/*γ* = 10 (days), respectively (4,5). In addition, we assumed that *S* + *E* + *I* + *R* = 1 so that each population implies the proportion to the total population. To estimate the infection rate *β*, we assumed that the daily number of newly reported cases is given by *Y* = *pIN* and fitted it to the data (2) to find the best *β* that minimizes the least-squares error, where *p* denotes the detection rate and *N* denotes the population in Tokyo. We assumed that *N* = 1.39 × 10^7^ (6), and we also estimated *p* by using the observational fact that antibody prevalence in Tokyo was 0.1% in June 2020 (7) and 1.35% in December 2020 (8). More precisely, we estimated *p* so that the removed population *R* in June and December 2020 is close to 0.001 and 0.0135, respectively, under the assumption that the antibody prevalence can be approximated by *R*. As stated above, the estimation was updated per week, and hence, we obtained the infection rate *β* in each week from January 23, 2020 to April 16, 2021. To predict the epidemic curve after April 17, 2021, we calculated the average of the infection rates in the last three weeks and assumed that it will not change. To discuss the effect of the vaccination, we introduced the vaccination rate *v* and changed the equations of *S* and *R* as *S*′ = −*βSI* − *vS* and *R*′ = *γI* + *vS*, respectively.

Figure 1 depicts the curve of new cases (1A, upper figure) and antibody prevalence (1B, lower figure). Our curve predicts that the maximum daily number of the infected cases in Tokyo in August 2021 would be 7,991 if the current pace of vaccinations (1/1,000 per day of the adult population, about 10,000 two-shot complete vaccinations). This daily number is greater than the highest daily cases (2,447) recorded on January 7 (9). During that month in Tokyo, the healthcare system was overwhelmed.

**Figure 1.**
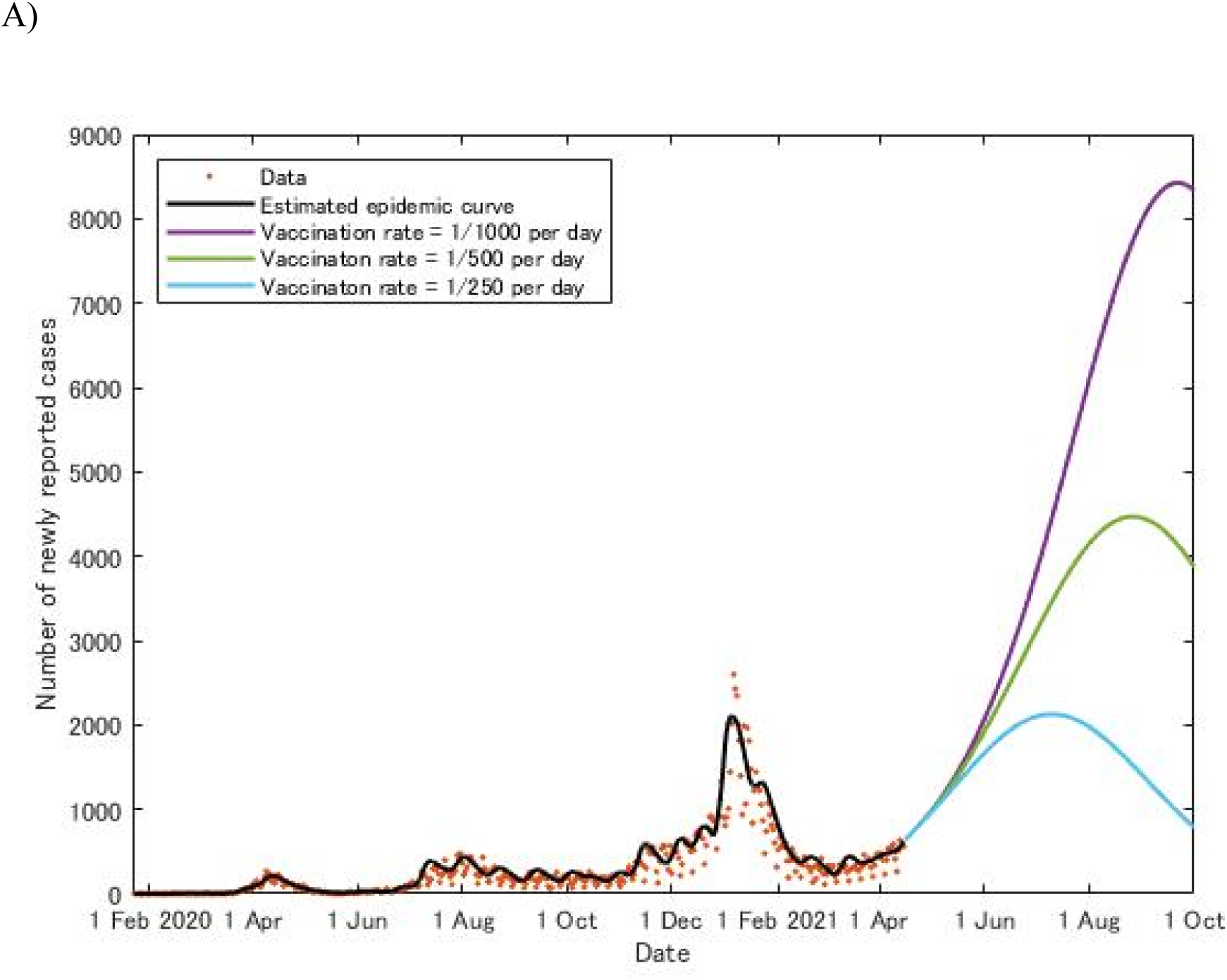
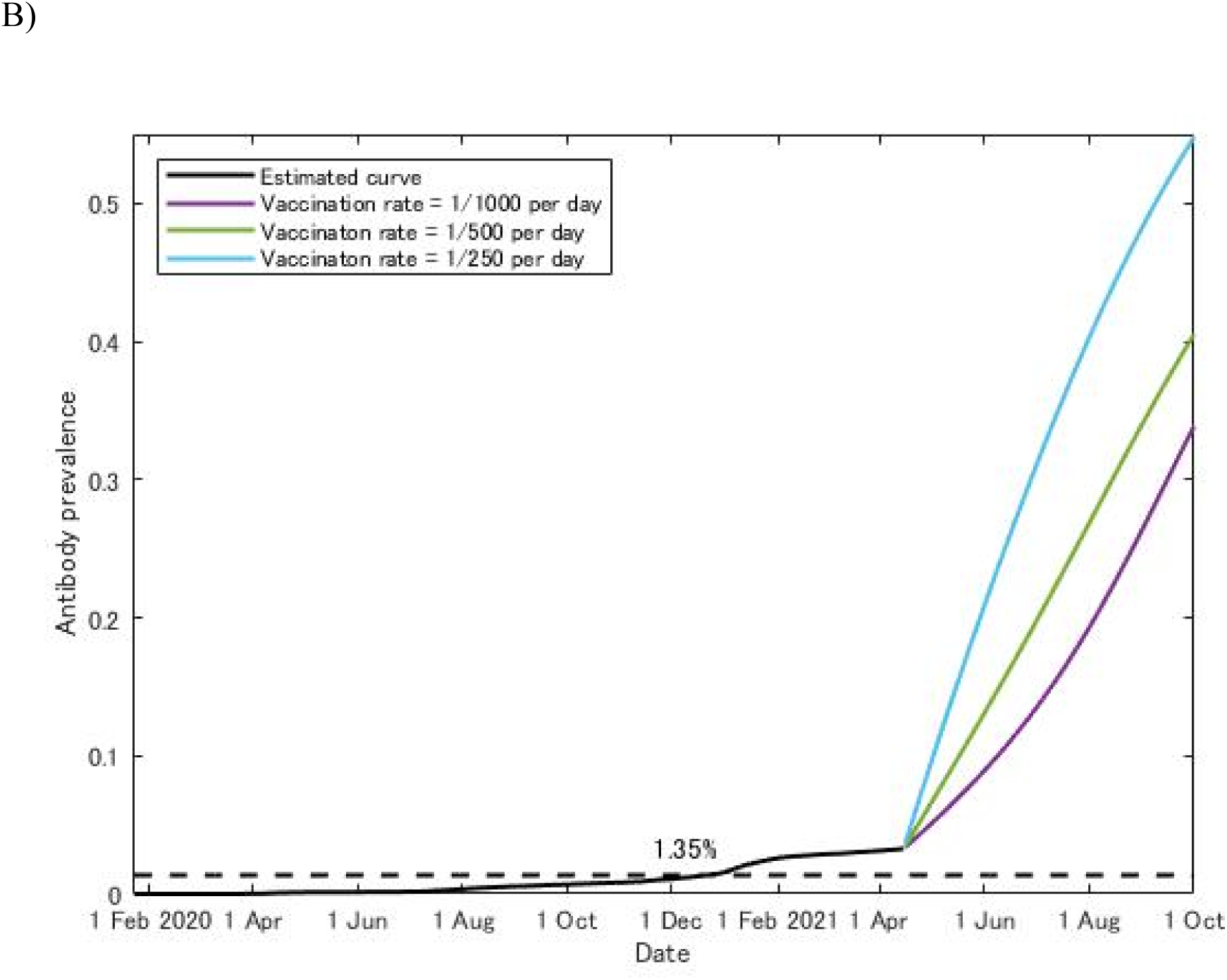
Legend A) The number of newly reported or predicted daily cases of COVID-19 in Tokyo B) Antibody prevalence for COVID-19 in Tokyo

However, if the rollout pace could be doubled (1/500 per day), the peak daily new cases would be 4,470 in August. If it could be quadrupled (1/250 per day), the peak would be noted at 2,128 in July and the highest number in August would be 1,977. In these accelerated pace scenarios, subsequent cases would be decreasing thereafter (Figure 1A). The limitation of our analysis included unclear effects from possible public health intervention such as strict lockdown as well as from higher transmission of variants. Nevertheless, based on our predictive analysis, the vaccination speed in Tokyo should be rapidly increased to mitigate a COVID-19 surge to prevent overwhelmed local healthcare system and less availability of healthcare workers during the time of the Games. This might be able to lower risk of infection among athletes and all the people involved in the Games. If vaccine rollout could not be enhanced before the Games, the cancellation might be an acceptable decision. After all, health is the most precious to our Olympians.

## Data Availability

All data referred to in the manuscript are available upon request.

There are no conflicts of interest to declare.

No funding.

Ethics committee approval: N.A.

